# The relative contribution of COVID-19 infection versus COVID-19 related occupational stressors to insomnia in healthcare workers

**DOI:** 10.1101/2022.10.27.22281582

**Authors:** Rebecca C. Hendrickson, Catherine A. McCall, Aaron F. Rosser, Kathleen F. Pagulayan, Bernard P. Chang, Ellen D. Sano, Ronald G. Thomas, Murray A. Raskind

**Author notes:** Corresponding Author; 1660 S Columbian Way, S116 MRECC, Seattle, WA 98108, USA.

## Abstract

**Objective/Background:** Healthcare workers have experienced high rates of psychiatric symptom burden and occupational attrition during the COVID-19 pandemic. Identifying contributory factors can inform prevention and mitigation measures. Here, we explore the potential contributions of occupational stressors vs COVID-19 infection to insomnia symptoms in US healthcare workers.

**Patients/Methods:** An online self-report survey was collected between September 2020 and July 2022 from N=594 US healthcare workers, with longitudinal follow-up up to 9 months. Assessments included the Insomnia Severity Index (ISI), the PTSD Checklist for DSM-5 (PCL-5), and a 13-item scale assessing COVID-19 related occupational stressors.

**Results:** Insomnia was common (45% of participants reported at least moderate and 9.2% reported severe symptoms at one or more timepoint) and significantly associated with difficulty completing work-related tasks, increased likelihood of occupational attrition, and thoughts of suicide or self-harm (all p<.0001). In multivariable regression with age, gender, and family COVID-19 history as covariates, past two-week COVID-related occupational stressors, peak COVID-related occupational stressors, and personal history of COVID-19 infection were all significantly related to past two-week ISI scores (β=1.7±0.14SE, β=0.08±0.03, and β=0.69±0.22 respectively). Although similar results were found for the PCL-5, when ISI and PCL-5 items were separated by factor, COVID-19 infection was significantly related only to the factor consisting of sleep-related items.

**Conclusions:** Both recent occupational stress and personal history of COVID-19 infection were significantly associated with insomnia in healthcare workers. These results suggest that both addressing occupational stressors and reducing rates of COVID-19 infection are important to protect healthcare workers and the healthcare workforce.

## Introduction

Healthcare workers during the COVID-19 pandemic have experienced significant and sustained elevations in psychiatric distress^1–3^, as well as increased frequency of suicidal ideation, worsened occupational functioning, and increased planned or actual workplace attrition. Identifying specific causes and mitigation strategies for these adverse outcomes is an important part of building resilience for the healthcare system during this and future pandemics or other stressors.

We and others have found significant dose-response relationships between the intensity of COVID-19 related occupational stressors experienced and negative psychiatric and occupational outcomes in healthcare workers, supporting a causal role of stressful and demoralizing experiences for healthcare workers working during the pandemic on psychiatric and occupational outcomes^1,4^. Alternatively, acute infection with COVID-19 has itself been found to be associated with persistent increases in psychiatric distress and worsened functional outcomes in a wide variety of populations^5^, including healthcare workers^6^. As high rates of exposure to COVID-19 related occupational stressors and elevated risk of COVID-19 infection have often gone hand in hand for healthcare workers, it has been challenging to assess the relative contributions of COVID-19 infection and occupational stress and trauma for healthcare worker health and occupational outcomes.

The potential for both occupational stressors and persistent effects of COVID-19 infection to drive symptom burden and adverse outcomes is particularly significant for insomnia symptoms. Although an increase in primarily subclinical insomnia symptoms has been observed in the general population during the COVID-19 pandemic^7^, significantly higher rates of pandemic-associated insomnia have been reported in healthcare workers^2,8^. Insomnia burden has been associated with a range of adverse personal and occupational outcomes, including increases in medical morbidity, suicidality, professional burnout, and the frequency of both errors and absences in the workplace^9–11^. As a result, identifying the factors driving increased symptoms of insomnia in healthcare workers is an important priority. Observational studies have identified increased workload or work-related stressors and anxiety about occupational exposure risk as significant contributors to the elevated rates of insomnia in healthcare workers^1,4^. However, persistent increases in physiologic arousal, including insomnia, have also been identified as some of the most common prolonged sequelae of COVID-19 infection^5,12^. An improved understanding of the relative contribution of occupational stressors versus a personal history of COVID-19 infection to the total burden of insomnia symptoms in healthcare workers working during the COVID-19 pandemic would help to guide workplace interventions to protect healthcare workers and our healthcare system.

Here, we draw on longitudinal assessments of exposure to COVID-19 related occupational stressors, COVID-19 infection history, and insomnia and psychiatric symptoms to examine the simultaneous contributions of both COVID-19 related occupational stressors and personal history of COVID-19 infection to insomnia symptoms in healthcare workers, and the impact of those symptoms on suicidality and adverse occupational outcomes.

## Methods

### 1. Participants

As described in Hendrickson *et al*.^1^, results were obtained from a national internet-based survey of US healthcare workers and first responders; the present work includes responses from 594 US health care workers (including 83 physicians and 388 nurses) collected between September 15, 2020 and July 17, 2022. Longitudinal follow-up was available for 350 participants, with a mean follow-up duration of 89 days (max: 291 days). The study was approved by the VA Puget Sound Health Care System Human Subjects Committee. Prior to enrollment, all participants were provided an information statement that detailed the purpose, risks, benefits, and alternatives to participation.

### 2. Measures

Insomnia was assessed with the Insomnia Severity Index (ISI)^13^, PTSD symptoms with the PTSD Checklist for DSM-5 (PCL-5)^14^, and thoughts of suicide or self-harm by item 9 of the Patient Health Questionnaire 9-item (PHQ-9)^15^. COVID-19 related occupational stressors were quantified using a previously published 13-item scale^1^. Functional impairment was assessed using the two work-related items from the PROMIS Short Form v2.0 Ability to Participate in Social Roles and Activities 8a measure^16^, modified to focus on occupational work^1^. Likelihood of leaving one’s current profession was assessed with two custom items^1^.

### 3. Data analysis

Data were analyzed using R and RStudio. Figures were created using *ggplot2*^17^ and *jtools*^18^. For all analyses, we selected a p-value of <.05 to determine statistical significance. Participants with missing data were excluded from analyses using items or measures with missing data. Multivariable linear regression models were implemented using *nlme*^19^, with a corAR1 correlation structure to account for repeated measures within subjects. Factor analysis was implemented using *psych* and *psychTools*^20^ using a varimax rotation.

## Results

Insomnia was common in the sample, with a mean ISI at baseline of 12.3±5.7 (36% with moderate or greater insomnia, 5.1% with severe insomnia; longitudinally, 45% of participants reported an ISI in the moderate or greater range at some point, and 9.2% reported an ISI in the severe range at some point; see table). ISI total score was significantly and positively associated with self-reported difficulty with completing work-related tasks, decreased likelihood of remaining in one’s current field, and thoughts of suicide or self-harm (all p<.00001, adjusted for age and gender; see supplementary material for additional models and details).

**Table:**

|  | Baseline Assessments<br>(N=594) | Maximal value for each<br>participant across<br>longitudinal assessments |
| --- | --- | --- |
| Age (years, mean±sd) | 42.9±10.9 |  |
| % female | 71.7% |  |
| % physician | 14.0% |  |
| % nurse | 65.3% |  |
| History of COVID-19<br>infection | 34.7% | 41.2% |
| COVID-19 related<br>occupational stress total<br>score (mean±sd) | 14.9±8.7 | 18.6±9.1 |
| ISI total score (mean±sd) | 12.3±5.76 | 13.6±5.9 |
| % moderate or greater<br>(≥15) | 35.9% | 45.4% |
| % severe (≥22) | 5.2% | 9.2% |
| PCL-5 total score<br>(mean±sd) | 28.8±18.4 | 31.8±19.1 |
| % clinical range (≥31) | 40.1% | 46.5% |
| % with past 2-weeks<br>thoughts of suicide or self-harm | 14.8% | 18.1% |

In a multivariable regression model with age, gender and history of COVID-19 infection in a family member as covariates, past two-week intensity of COVID-19 related occupational stress, peak intensity of COVID-19 related occupational stress, and personal history of COVID-19 infection were all independent, significant predictors of past two-week insomnia symptoms. While past two-week COVID-related occupational stress was the factor most strongly associated with insomnia symptoms (β=1.7±0.14SE, p<1e-4), personal history of COVID-19 infection had a substantially stronger association with insomnia symptoms than peak COVID-19 related occupational stress (β=0.69±0.22 vs β=0.08±0.03, both p<.01).

To explore the specificity of the association, we explored the relationship of the same predictor variables to PTSD symptoms as quantified by PCL-5 total score (fig 1A). As with insomnia, past two-week COVID-related occupational stressors were the strongest predictor of PTSD symptoms, followed by personal history of COVID-19 infection and then peak COVID-related occupational stressors. Because the PCL-5 contains items related to sleep, however, we also carried out a factor analysis of the pooled ISI and PCL-5 questions based on responses in our data set. This analysis identified 3 stable factors (fig 1B): factor 1 emphasized items related to affect and arousal; factor 2 emphasized items specific to PTSD; and factor 3 emphasized items related to insomnia. Of these factors, personal history of COVID-19 infection was a significant predictor only of the Insomnia factor (Fig 1A), and only personal history of COVID-19 infection and past two-weeks COVID-19 related occupational stressors were significant predictors of this factor, with peak COVID-related occupational stressors no longer significant as a predictor.

**Figure 1:**
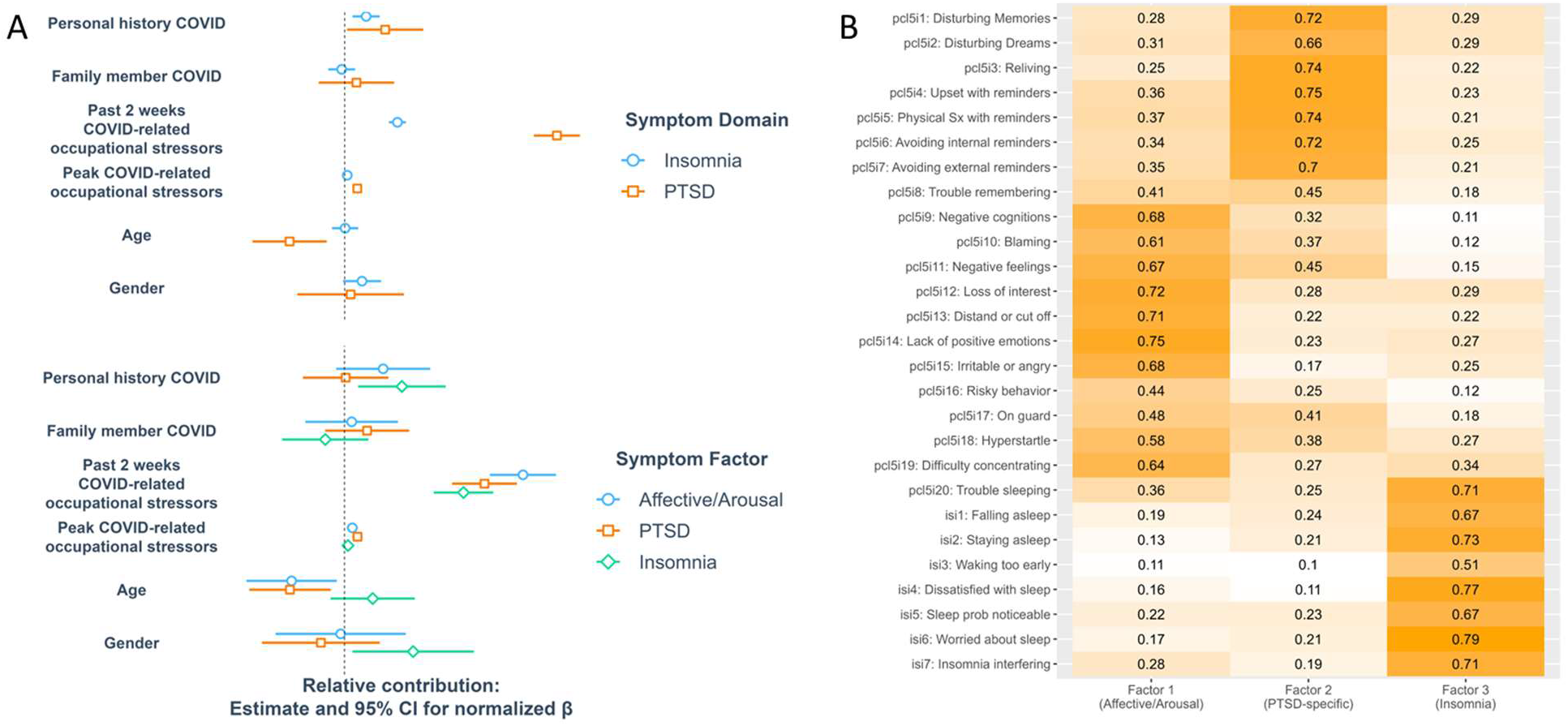
Relationship of personal history of COVID-19 infection and exposure to COVID-19 related occupational stressors to (A, top) insomnia and PTSD symptoms, and (A, bottom) 3 factors of ISI and PCL-5 pooled items, emphasizing affective/arousal items, PTSD-specific items, and insomnia items (B). Panel (A) represents the results of independent multivariable regression models relating past two-week total score and peak total score on COVID-related occupational stressor scale, personal history of COVID-19 infection, and covariates of age and gender to total scores on the ISI and PCL-5 or factor totals; y-axis shows point estimate and SE of β using normalized predictor variables.

## Discussion

Consistent with previous literature^2,8^, insomnia was prevalent in healthcare workers working during the COVID-19 pandemic, with nearly half of participants reporting at least moderate insomnia at some point during their participation and nearly 10% reporting severe insomnia during this period, and was associated with decreased self-reported occupational functioning, increased self-reported likelihood of leaving the field, and thoughts of suicide or self-harm.

We found that both COVID-19 related occupational stressors over the past two weeks and any prior history of COVID-19 infection were significantly associated with past two-weeks insomnia symptoms, even accounting for prior (peak) COVID-19 related occupational stressor burden. Using a factor analysis of pooled items from both PTSD- and insomnia-focused symptom self-report scales, we found a specific association of prior COVID-19 infection and current insomnia symptoms, with no effect seen for PTSD-specific symptoms. This divergence is potentially consistent with insomnia symptoms resulting from a persistent biologic effect of COVID-19 infection, as has been suggested by recent epidemiologic findings^12^.

These results have implications for both provider well-being/health and for patient care. The high prevalence of sleep disturbances in our study and their strong association with measures of adverse both personal and occupational outcomes underscore the importance for individuals working in healthcare and for the resilience of our healthcare system of addressing causes of insomnia in the healthcare workforce. In addition, to the extent persistent insomnia after COVID-19 acute infection is a significant cause of these high levels of insomnia and the associated adverse outcomes, rather than COVID-19 related occupational stressors alone, it is likely that these findings may have relevance to individuals outside the healthcare workforce who experience COVID-19 infection, as well.

One limitation of the present work is that we did not collect information about the timing of COVID-19 infection in those whose infection occurred prior to their baseline assessment. Thus, we are unable to quantify the duration of this effect after infection. Nonetheless, these results suggest that persistent symptoms following COVID-19 infection are a significant contributor to the overall prevalence and severity of insomnia in healthcare workers, even when both ongoing and prior stress related to working during the pandemic are taken into account. These findings highlight the importance of reducing not only COVID-19 related occupational stressors, but also the risk of acute COVID-19 infection itself, in interventions seeking to protect the mental and physical health of healthcare workers and the resilience of our healthcare system.

## Data Availability

Deidentified data sets may be requested by contacting the corresponding author.

## Contributors

The authors would like to thank the many participants who generously shared their time and experiences with us.

## Contributions of authors

RCH designed the study, with input from BPC, CAM, and MAR. RCH supervised the implementation of the study and carried out the analysis of all results with consultation by RGT. RCH wrote the initial draft of the manuscript, with contributions from AFR. All authors provided input into the interpretation of the analysis and the drafting of the manuscript.

## Data sharing

Deidentified data sets may be requested by contacting the corresponding author.

