## Supplemental Materials for "The relative contribution of COVID-19 infection versus COVID-19 related occupational stressors to insomnia in healthcare workers"

Supplemental Materials: Details of statistical regression models

(1) Models addressing relationship of insomnia as measured by the ISI to various outcomes, as described in text:

a) Model relating sleep to occupational functioning:

Generalized least squares fit by REML

Model: work ~ isi_total + age + gender

| AIC | BIC | logLik |
|---|---|---|
| 6160.261 | 6192.799 | -3074.131 |

Correlation Structure: AR(1)

Formula: ~1 | PtID

Parameter estimate(s):

| Phi |
|---|
| **0.6263705** |

Coefficients:

| | Value | Std. Error | t-value | p-value |
|---|---|---|---|---|
| **(Intercept)** | 2.2535563 | 0.12275909 | 28.359910 | 0.0000 |
| **ISI Total** | 0.0909617 | 0.00867815 | 10.481693 | 0.0000 |
| **age** | -0.0961472 | 0.07137116 | -1.347144 | 0.1781 |
| **gender** | 0.1531653 | 0.10542437 | 1.452846 | 0.1465 |

| | | |
|---|---|---|
| **Residual Standard Error** | 1.819436 | |
| **Degrees of Freedom** | 1678 total | 1674 residual |

b) Model relating sleep to self-reported likelihood of leaving current field:

Generalized least squares fit by REML

Model: infield ~ isi_total + age + gender

| AIC | BIC | logLik |
|---|---|---|
| 5527.824 | 5560.351 | -2757.912 |

Correlation Structure: AR(1)

Formula: ~1 | PtID

Parameter estimate(s):

| Phi |
|---|
| **0.7662417** |

Coefficients:

| | Value | Std. Error | t-value | p-value |
|---|---|---|---|---|
| **(Intercept)** | 3.253587 | 0.11472488 | 28.359910 | 0.0000 |
| **ISI Total** | 0.050418 | 0.00734686 | 6.862578 | 0.0000 |
| **age** | 0.211740 | 0.07631342 | 2.774610 | 0.0056 |
| **gender** | 0.085483 | 0.11137435 | 0.767531 | 0.4429 |

| | | |
|---|---|---|
| **Residual Standard Error** | 1.750745 | |
| **Degrees of Freedom** | 1675 total | 1671 residual |

a) Model relating sleep to thoughts of suicide, self-harm, or being better off dead:

Generalized least squares fit by REML

Model: phq9i9 ~ isi_total + age + gender

| AIC | BIC | logLik |
|---|---|---|
| 1060.662 | 1088.358 | -524.331 |

Correlation Structure: AR(1)

Formula: ~1 | PtID

Parameter estimate(s):

| Phi |
|---|
| **0.5447143** |

Coefficients:

| | Value | Std. Error | t-value | p-value |
|---|---|---|---|---|
| **(Intercept)** | - 0.10044693 | 0.04722915 | - 2.126799 | 0.0338 |
| **ISI Total** | 0.02447273 | 0.00349304 | 7.006141 | 0.0000 |
| **age** | - 0.02131648 | 0.02412003 | - 0.883767 | 0.3771 |
| **gender** | 0.03231543 | 0.03481727 | 0.928144 | 0.3536 |

| | | |
|---|---|---|
| **Residual Standard Error** | 0.5206405 | |
| **Degrees of Freedom** | 751 total | 747 residual |

(2) Core model addressing predictors of ISI from text:

Generalized least squares fit by REML

Model: ISI total score ~ Personal history of COVID-19 infection + Family member with COVID-19 infection + Past 2 weeks COVID-related occupational stressors + Peak COVID-related occupational stressors + age + gender

| AIC | BIC | logLik |
|---|---|---|
| 9392.458 | 9441.313 | -4687.229 |

Correlation Structure: AR(1)

Formula: ~1 | PtID

Parameter estimate(s):

| Phi |
|---|
| **0.7159753** |

Coefficients:

| | Value | Std. Error | t-value | p-value |
|---|---|---|---|---|
| **(Intercept)** | 9.726505 | 0.5120368 | 18.995716 | 0.0000 |
| **Personal history of COVID-19 infection** | 0.692786 | 0.2236221 | 3.098021 | 0.0020 |
| **Family member with COVID-19 infection** | -0.096676 | 0.2210195 | -0.437409 | 0.6619 |
| **Past 2 weeks COVID-related occupational stressors** | 1.704757 | 0.1402930 | 12.151402 | 0.0000 |
| **Peak COVID-related occupational stressors** | 0.082858 | 0.0256810 | 3.226414 | 0.0013 |
| **Age** | 0.014379 | 0.2163650 | 0.066459 | 0.9470 |
| **Gender** | 0.559322 | 0.3131433 | 1.786152 | 0.0743 |

Correlation:

| | (Intr) | PERSONAL HISTORY OF COVID-19 INFECTION | FAMILY MEMBER WITH COVID-19 INFECTION | Past 2 weeks COVID-related | PEAK COVID-19 RELATED OCCUPATIONAL STRESSORS | age |
|---|---|---|---|---|---|---|

| | | | | occupational stressors | | |
|---|---|---|---|---|---|---|
| **Personal history of COVID-19 infection** | 0.170 | | | | | |
| **Family member with COVID-19 infection** | 0.041 | -0.312 | | | | |
| **Past 2 weeks COVID-related occupational stressors** | 0.328 | -0.014 | -0.036 | | | |
| **Peak COVID-related occupational stressors** | -0.902 | -0.186 | -0.027 | -0.381 | | |
| **age** | -0.108 | 0.064 | -0.103 | 0.041 | 0.120 | |
| **gender** | -0.069 | 0.025 | 0.000 | 0.015 | -0.096 | -0.007 |

Standardized residuals:

| Min | Q1 | Med | Q3 | Max |
|---|---|---|---|---|
| -2.88856050 | -0.67696599 | 0.03740746 | 0.67985578 | 2.97731249 |

| | | |
|---|---|---|
| **Residual Standard Error** | 5.090767 | |
| **Degrees of Freedom** | 1690 total | 1683 residual |

(3) Models for Fig 1A top

> summary(fitisi)

Generalized least squares fit by REML

Model: ISI total score ~ Personal history of COVID-19 infection + Family member with COVID-19 infection + Past 2 weeks COVID-related occupational stressors + Peak COVID-related occupational stressors + age + gender

| AIC | BIC | logLik |
|---|---|---|
| 9392.458 | 9441.313 | -4687.229 |

Correlation Structure: AR(1)

Formula: ~1 | PtID

Parameter estimate(s):

| Phi |
|---|
| **0.7159753** |

Coefficients:

| | Value | Std.Error | t-value | p-value |
|---|---|---|---|---|
| **(Intercept)** | 9.726505 | 0.5120368 | 18.995716 | 0.0000 |
| **Personal history of COVID-19 infection** | 0.692786 | 0.2236221 | 3.098021 | 0.0020 |
| **Family member with COVID-19 infection** | -0.096676 | 0.2210195 | -0.437409 | 0.6619 |
| **Past 2 weeks COVID-related occupational stressors** | 1.704757 | 0.1402930 | 12.151402 | 0.0000 |
| **Peak COVID-related occupational stressors** | 0.082858 | 0.0256810 | 3.226414 | 0.0013 |
| **age** | 0.014379 | 0.2163650 | 0.066459 | 0.9470 |
| **gender** | 0.559322 | 0.3131433 | 1.786152 | 0.0743 |

Correlation:

| | (Intr) | PERSONAL HISTORY OF COVID-19 INFECTION | FAMILY MEMBER WITH COVID-19 INFECTION | Past 2 weeks COVID-related occupational stressors | PEAK COVID-19 RELATED OCCUPATIONAL STRESSORS | age |
|---|---|---|---|---|---|---|
| **Personal history of COVID-19 infection** | 0.170 | | | | | |
| **Family member with COVID-19 infection** | 0.041 | -0.312 | | | | |
| **Past 2 weeks COVID-related occupational stressors** | 0.328 | -0.014 | -0.036 | | | |
| **Peak COVID-related occupational stressors** | -0.902 | -0.186 | -0.027 | -0.381 | | |

| | | | | | | |
|---|---|---|---|---|---|---|
| **age** | -0.108 | 0.064 | -0.103 | 0.041 | 0.120 | |
| **gender** | -0.069 | 0.025 | 0.000 | 0.015 | -0.096 | -0.007 |

Standardized residuals:

| Min | Q1 | Med | Q3 | Max |
|---|---|---|---|---|
| -2.88856050 | -0.67696599 | 0.03740746 | 0.67985578 | 2.97731249 |

| **Residual standard error** | 5.090767 | |
|---|---|---|
| **Degrees of freedom** | 1690 total | 1683 residual |

> summary(fitpcl)

Generalized least squares fit by REML

Model: pcl5total ~ Personal history of COVID-19 infection + Family member with COVID-19 infection + Past 2 weeks COVID-related occupational stressors + Peak COVID-related occupational stressors + age + gender

Data: data2use

| AIC | BIC | logLik |
|---|---|---|
| 12382.31 | 12430.92 | -6182.157 |

Correlation Structure: AR(1)

Formula: ~1 | PtID

Parameter estimate(s):

| Phi |
|---|
| **0.738682** |

Coefficients:

| | Value | Std. Error | t-value | p-value |
|---|---|---|---|---|
| **Intercept** | 17.113036 | 1.4329872 | 11.942211 | 0.0000 |
| **Personal history of COVID-19 infection** | 1.307692 | 0.6287211 | 2.079924 | 0.0377 |
| **Family member with COVID-19 infection** | 0.379777 | 0.6227244 | 0.609864 | 0.5420 |
| **Past 2 weeks COVID-related occupational stressors** | 6.857538 | 0.3801570 | 18.038703 | 0.0000 |

| | | | | |
|---|---|---|---|---|
| **Peak COVID-related occupational stressors** | 0.407807 | 0.0716705 | 5.690021 | 0.0000 |
| **age** | -1.779545 | 0.6111902 | -2.911606 | 0.0036 |
| **gender** | 0.192778 | 0.8780037 | 0.219563 | 0.8262 |

Correlation:

| | (Intr) | PERSONAL HISTORY OF COVID-19 INFECTION | FAMILY MEMBER WITH COVID-19 INFECTION | Past 2 weeks COVID-related occupational stressors | PEAK COVID-19 RELATED OCCUPATIONAL STRESSORS | age |
|---|---|---|---|---|---|---|
| **Personal history of COVID-19 infection** | 0.169 | | | | | |
| **Family member with COVID-19 infection** | 0.046 | -0.308 | | | | |
| **Past 2 weeks COVID-related occupational stressors** | 0.315 | -0.012 | -0.031 | | | |
| **Peak COVID-related occupational stressors** | -0.900 | -0.186 | -0.031 | -0.370 | | |
| **age** | -0.106 | 0.061 | -0.105 | 0.036 | 0.119 | |
| **gender** | -0.068 | 0.024 | 0.001 | 0.014 | -0.096 | -0.012 |

Standardized residuals:

| Min | Q1 | Med | Q3 | Max |
|---|---|---|---|---|
| -2.4462126 | -0.6925527 | -0.1903986 | 0.5415783 | 4.3297910 |

| | | |
|---|---|---|
| **Residual Standard Error** | 14.05411 | |
| **Degrees of Freedom** | 1644 total | 1637 residual |

(4) Models for Fig1A bottom

> summary(fitF1)

Generalized least squares fit by REML

Model: mhsmall3f_1 ~ Personal history of COVID-19 infection + Family member with COVID-19 infection + Past 2 weeks COVID-related occupational stressors + Peak COVID-related occupational stressors + age + gender

Data: data2use

| AIC | BIC | logLik |
|---|---|---|
| 4140.04 | 4188.602 | -2061.02 |

Correlation Structure: AR(1)

Formula: ~1 | PtID

Parameter estimate(s):

| Phi |
|---|
| **0.6597996** |

Coefficients:

| | Value | Std. Error | t-value | p-value |
|---|---|---|---|---|
| **Intercept** | 0.8688129 | 0.10431685 | 8.328596 | 0.0000 |
| **Personal history of COVID-19 infection** | 0.0724225 | 0.04499485 | 1.609573 | 0.1077 |
| **Family member with COVID-19 infection** | 0.0134383 | 0.04430643 | 0.303303 | 0.7617 |
| **Past 2 weeks COVID-related occupational stressors** | 0.3346461 | 0.03159189 | 10.592786 | 0.0000 |
| **Peak COVID-related occupational stressors** | 0.0146770 | 0.00525709 | 2.791850 | 0.0053 |
| **age** | -0.0992555 | 0.04316311 | -2.299545 | 0.0216 |
| **gender** | -0.0071763 | 0.06226016 | -0.115263 | 0.9083 |

Correlation:

| | (Intr) | PERSONAL HISTORY OF COVID-19 INFECTION | FAMILY MEMBER WITH COVID-19 INFECTION | Past 2 weeks COVID-related occupational stressors | PEAK COVID-19 RELATED OCCUPATIONAL STRESSORS | age |
|---|---|---|---|---|---|---|
| **Personal history of COVID-19 infection** | 0.177 | | | | | |
| **Family member with COVID-19 infection** | 0.037 | -0.306 | | | | |
| **Past 2 weeks COVID-related occupational stressors** | 0.367 | -0.014 | -0.039 | | | |
| **Peak COVID-related occupational stressors** | -0.906 | -0.189 | -0.020 | -0.417 | | |
| **age** | -0.106 | 0.064 | -0.112 | 0.043 | 0.111 | |
| **gender** | -0.062 | 0.030 | 0.000 | 0.019 | -0.099 | 0.004 |

Standardized residuals:

| Min | Q1 | Med | Q3 | Max |
|---|---|---|---|---|
| -3.1839267 | -0.7271638 | -0.1886885 | 0.5096133 | 3.7781573 |

| | | |
|---|---|---|
| **Residual Standard Error** | 1.049544 | |
| **Degrees of Freedom** | 1636 total | 1629 residual |

> summary(fitF2)

Generalized least squares fit by REML

Model: mhsmall3f_2 ~ Personal history of COVID-19 infection + Family member with COVID-19 infection + Past 2 weeks COVID-related occupational stressors + Peak COVID-related occupational stressors + age + gender

Data: data2use

| AIC | BIC | logLik |
|---|---|---|
| **4051.473** | 4100.035 | -2016.737 |

Correlation Structure: AR(1)

Formula: ~1 | PtID

Parameter estimate(s):

| Phi |
|---|
| **0.6101749** |

Coefficients:

| | Value | Std. Error | t-value | p-value |
|---|---|---|---|---|
| **Intercept** | 0.03333596 | 0.09584669 | 0.347805 | 0.7280 |
| **Personal history of COVID-19 infection** | 0.00219464 | 0.04090616 | 0.053651 | 0.9572 |
| **Family member with COVID-19 infection** | 0.04240595 | 0.04014654 | 1.056279 | 0.2910 |
| **Past 2 weeks COVID-related occupational stressors** | 0.26250679 | 0.03102805 | 8.460306 | 0.0000 |
| **Peak COVID-related occupational stressors** | 0.02437243 | 0.00484542 | 5.029998 | 0.0000 |
| **age** | -0.10231473 | 0.03897661 | -2.625029 | 0.0087 |
| **gender** | -0.04428009 | 0.05635074 | -0.785794 | 0.4321 |

Correlation:

| | (Intr) | PERSONAL HISTORY OF COVID-19 INFECTION | FAMILY MEMBER WITH COVID-19 INFECTION | Past 2 weeks COVID-related occupational stressors | PEAK COVID-19 RELATED OCCUPATIONAL STRESSORS | age |
|---|---|---|---|---|---|---|
| **Personal history of COVID-19 infection** | 0.181 | | | | | |
| **Family member with COVID-19 infection** | 0.032 | -0.308 | | | | |
| **Past 2 weeks COVID-related occupational stressors** | 0.396 | -0.015 | -0.044 | | | |
| **Peak COVID-related** | -0.909 | -0.191 | -0.014 | -0.443 | | |

| | | | | | | |
|---|---|---|---|---|---|---|
| **occupational stressors** | | | | | | |
| **age** | -0.105 | 0.065 | -0.116 | 0.047 | 0.106 | |
| **gender** | -0.058 | 0.034 | -0.002 | 0.022 | -0.101 | 0.013 |

Standardized residuals:

| Min | Q1 | Med | Q3 | Max |
|---|---|---|---|---|
| -3.4511443 | -0.6584667 | -0.1186989 | 0.5706452 | 4.0646472 |

| **Residual standard error:** | 0.9810603 | |
|---|---|---|
| **Degrees of freedom:** | 1636 total | 1629 residual |

> summary(fitF3)

Generalized least squares fit by REML

Model: mhsmall3f_3 ~ Personal history of COVID-19 infection + Family member with COVID-19 infection + Past 2 weeks COVID-related occupational stressors + Peak COVID-related occupational stressors + age + gender

Data: data2use

| AIC | BIC | logLik |
|---|---|---|
| 3812.102 | 3860.664 | -1897.051 |

Correlation Structure: AR(1)

Formula: ~1 | PtID

Parameter estimate(s):

| Phi |
|---|
| **0.6789277** |

Coefficients:

| | Value | Std.Error | t-value | p-value |
|---|---|---|---|---|
| **(Intercept)** | 1.5813248 | 0.09662911 | 16.364891 | 0.0000 |
| **Personal history of COVID-19 infection** | 0.1075206 | 0.04185497 | 2.568885 | 0.0103 |
| **Family member with COVID-19 infection** | -0.0359708 | 0.04126998 | -0.871597 | 0.3836 |
| **Past 2 weeks COVID-related occupational stressors** | 0.2230959 | 0.02843435 | 7.846000 | 0.0000 |

| | | | | |
|---|---|---|---|---|
| **Peak COVID-related occupational stressors** | 0.0065025 | 0.00486237 | 1.337318 | 0.1813 |
| **age** | 0.0528811 | 0.04026294 | 1.313394 | 0.1892 |
| **gender** | 0.1286761 | 0.05802223 | 2.217704 | 0.0267 |

Correlation:

| | **(Intr)** | **PERSONAL HISTORY OF COVID-19 INFECTION** | **FAMILY MEMBER WITH COVID-19 INFECTION** | **Past 2 weeks COVID-related occupational stressors** | **PEAK COVID-19 RELATED OCCUPATIONAL STRESSORS** | **age** |
|---|---|---|---|---|---|---|
| **Personal history of COVID-19 infection** | 0.175 | | | | | |
| **Family member with COVID-19 infection** | 0.039 | -0.306 | | | | |
| **Past 2 weeks COVID-related occupational stressors** | 0.355 | -0.014 | -0.037 | | | |
| **Peak COVID-related occupational stressors** | -0.905 | -0.189 | -0.023 | -0.407 | | |
| **age** | -0.106 | 0.063 | -0.110 | 0.041 | 0.113 | |
| **gender** | -0.063 | 0.029 | 0.000 | 0.018 | -0.098 | 0.000 |

Standardized residuals:

| **Min** | **Q1** | **Med** | **Q3** | **Max** |
|---|---|---|---|---|
| -3.26387164 | -0.68447668 | 0.08933059 | 0.70115020 | 3.09688658 |

| | | |
|---|---|---|
| **Residual standard error** | 0.9659589 | |
| **Degrees of freedom** | 1636 total | 1629 residual |
